# Rare germline genetic variation in PAX8 transcription factor binding sites and susceptibility to epithelial ovarian cancer

**DOI:** 10.1101/2023.03.22.23287587

**Authors:** Suzana A. M. Ezquina, Michelle Jones, Ed Dicks, Amber de Vries, Pei-Chen Peng, Kate Lawrenson, Rosario I. Corona, Jonthan Tyrer, Dennis Hazelett, James Brenton, Antonis Antoniou, Simon A. Gayther, Paul D. P. Pharoah

**Affiliations:** Centre for Cancer Genetic Epidemiology, Department of Public Health and Primary Care, University of Cambridge, UK; Center for Bioinformatics and Functional Genomics, Department of Biomedical Sciences, Cedars-Sinai Medical Center, Los Angeles, USA; Department of Computational Biomedicine, Cedars-Sinai Medical Center, Los Angeles; Center for Bioinformatics and Functional Genomics, Department of Biomedical Sciences, Cedars-Sinai Medical Center, Los Angeles; Department of Human Genetics, University of California, Los Angeles, USA; Centre for Cancer Genetic Epidemiology, Department of Oncology, University of Cambridge, UK; Department of Computational Biomedicine, Cedars-Sinai Medical Center, Los Angeles, USA; Department of Oncology, University of Cambridge, UK

**Author notes:** Correspondence to Michelle Jones.

## Abstract

Common genetic variation throughout the genome together with rare coding variants identified to date explain about a half of the inherited genetic component of epithelial ovarian cancer risk. It is likely that rare variation in the non-coding genome will explain some of the unexplained heritability, but identifying such variants is challenging. The primary problem is lack of statistical power to identifying individual risk variants by association as power is a function of sample size, effect size and allele frequency. Power can be increased by using burden tests which test for association of carriers of any variant in a specified genomic region. This has the effect of increasing the putative effect allele frequency.

PAX8 is a transcription factor that plays a critical role in tumour progression, migration and invasion. Furthermore, regulatory elements proximal to target genes of PAX8 are enriched for common ovarian cancer risk variants. We hypothesised that rare variation in PAX8 binding sites are also associated with ovarian cancer risk, but unlikely to be associated with risk of breast, colorectal or endometrial cancer.

We have used publicly-available, whole-genome sequencing data from the UK 100,000 Genomes Project to evaluate the burden of rare variation in PAX8 binding sites across the genome. Data were available for 522 ovarian cancers, 2560 breast cancers, 2465 colorectal cancers and 729 endometrial cancers and 2253 non-cancer controls. Active binding sites were defined using data from multiple PAX8 and H3K27 ChIPseq experiments.

We found no association between the burden of rare variation in PAX8 binding sites (defined in several ways) and risk of ovarian, breast or endometrial cancer. An apparent association with colorectal cancer was likely to be a technical artefact as a similar association was also detected for rare variation in random regions of the genome.

Despite the null result this study provides a proof-of -principle for using burden testing to identify rare, non-coding germline genetic variation associated with disease. Larger sample sizes available from large-scale sequencing projects together with improved understanding of the function of the non-coding genome will increase the potential of similar studies in the future.

## Introduction

Genome-wide association studies (GWAS) conducted by the Ovarian Cancer Association Consortium (OCAC) identified 42 common epithelial ovarian cancer (EOC) susceptibility alleles^1^ and rare coding variants in at least 10 genes are known to be involved in susceptibility to epithelial ovarian cancer^2^. The high-penetrance genes *BRCA1*^3^ and *BRCA2*^4^ were identified by linkage studies in the 1990’s; protein truncating variants in these genes confer a substantial lifetime risk of EOC as well as breast cancer and other cancers^5,6^. Since then, truncating variants in *BRIP1, FANCM, PALB2, RAD51C* and *RAD51D* have been shown to confer more moderate risks by using candidate-gene case-control sequencing^7^. The uncommon and rare, high- and moderate penetrance alleles identified to date explain about one quarter of the inherited component of EOC susceptibility with another 5 percent explained by the known common risk alleles^2^. Genome-wide heritability analyses have estimated that the set of common variants that are tagged or captured by the standard genome-wide genotyping arrays explains about 40 percent of the familial aggregation – the so-called chip heritability^2^. The characteristics of the alleles that account for the remaining familial aggregation are not known; analyses of whole-genome data suggest that a substantial portion is explained by rare variants, particularly those in regions of low linkage disequilibrium ^8^.

The identification of rare genetic variants associated with disease susceptibility presents a considerable challenge. In principle it would be possible to sequence a very large number of cases and controls and to evaluate the disease associations on a variant-by-variant basis. This, in essence, has been the approach to common risk variant discovery in genome-wide association studies. However, the cost of sequencing tens of thousands of cases and controls remains prohibitively expensive. Even then the statistical power will be limited to detect the association on individual rare variants. Candidate gene sequencing was successful in identifying uncommon, loss-of-function variation associated with disease risk because it was been possible to apply gene-based burden tests in which any putative protein truncating variant in a gene is treated as equivalent and so the combined frequency of the variants is sufficient for reasonable sample sizes to have adequate power. The association of loss-of-function variants in *BRIP1, PALB2, RAD51C*, and *RAD51D* were identified using this approach.

Burden testing can also be applied to the non-coding genome. However, it is less clear what the unit for genomic analysis ought to be. One possibility is to use functional annotation to define non-contiguous genomic regions for burden testing. Such units could include all the binding sites for transcription factors of interest, regions of active chromatin in tissues of interest, regions of open chromatin in tissues of interest and regions that are frequently somatically mutated in ovarian cancer. The majority of known germline risk alleles associated with heritability of EOC risk is in non-protein coding regions, mostly located in active regulatory elements^9^. Putative regulatory elements proximal to target genes of critical transcription factors that are implicated in ovarian cancer or precursor cell biology are enriched for ovarian cancer risk SNPs, highlighting a central role for the transcription factor paired box 8 (PAX8)^10^. PAX8 is highly expressed in high-grade serous ovarian cancers and plays a critical role in tumour progression, migration and invasion^11^. PAX8 is also expressed in secretory epithelial cells of the fallopian tube, the precursor tissues of high-grade serous ovarian cancer ^12^. Thus, rare germline genetic variation in PAX8 bindings sites are candidate susceptibility alleles for EOC.

In addition to the PAX8 binding sites, we are also interested in active regions of the genome, where DNA is highly accessible, such as those identified by the histone H3K27 acetylation mark^13^. It has been shown that this important histone mark can distinguish between active and poised enhancer elements^14^, which tend to cluster near the genes they regulate. Genes proximal to H3K27ac marks also show higher expression levels^13^. Transcription factors such as PAX8 bind only to a subset of enhancers in a given tissue and H3K27ac marks can be used to select some of these enhancer locations. Previous analyses have shown that more than 80 percent of cell type specific regulatory elements lie in putative enhancers, which drive the cell type specific gene expression^2^.

The aim of this study was to use publicly available whole genome sequencing data from the UK 100,000 Genomes Project to evaluate the association between rare variation in active PAX8 binding sites and epithelial ovarian cancer risk, using a burden testing approach and a case-control study design. We also evaluated the association with breast, colorectal and endometrial cancer, with the goal of investigating the relevance of these target regions in other types of cancer.

## Methods

### Samples

The UK 100,000 Genomes Project { https://www.genomicsengland.co.uk/about-genomics-england/the-100000-genomes-project/} was established to sequence 100,000 genomes from around 85,000 NHS patients affected by a rare disease, or cancer. Recruitment of participants to the 100,000 Genomes Project was completed in 2018, with the 100,000th sequence achieved in December 2018. Germline whole genome sequencing has been carried out on 581 cases of EOC as part of the project (v15), from which 522 were included in the full analysis after filtering 59 outliers (see Methods -Sample quality control).

The majority of EOC was classified as high-grade serous carcinoma (n = 296); the number of cases of other histotypes are described in Table 1**Error! Reference source not found**.. We also analysed 2,984 cases of breast cancer, 2,696 cases of colorectal cancer and 836 cases of endometrial cancer, respectively.

**Table 1:** Number of epithelial ovarian cancer patients by histotype (total 522)

| Histotype | Number of individuals | Percent |
| --- | --- | --- |
| High grade serous* | 296 | 57% |
| Low grade serous | 25 | 5% |
| Clear cell | 33 | 6% |
| Endometrioid | 54 | 10% |
| Mucinous | 29 | 6% |
| Unknown | 72 | 14% |
| Other | 13 | 2% |

\* Includes those with carcinosarcoma and serous carcinoma of unspecified grade.

Unaffected controls have not been sequenced as part of the Project. However, many of the Project samples are from individuals with rare disorders that are extremely unlikely to share heritability with epithelial ovarian cancer and can be used as controls for the association analysis. We selected 2,750 patients as controls, from Genomics England main programme v6, with diseases such as myopathies and epilepsies, whose aetiology is multifactorial and unlikely to be related to cancer (Supplementary Table 1).

**Supplementary Table 1:** Diagnoses for 2,253 individuals selected as controls (497 were excluded as outliers)

| **Disease** | **Individuals** | **Disease** | **Individuals** |
| --- | --- | --- | --- |
| Amyotrophic lateral sclerosis | 23 | Epilepsy plus other features | 580 |
| Arthrogryposis | 40 | Epileptic encephalopathy | 112 |
| Brugada syndrome | 61 | Familial focal epilepsy | 21 |
| Complex Parkinsonism | 45 | Familial genetic generalised epilepsies | 61 |
| Congenital hearing impairment | 165 | Genetic epilepsies with febrile seizures | 8 |
| Congenital myopathy | 120 | Hereditary ataxia | 352 |
| Dilated cardiomyopathy | 115 | Hypertrophic cardiomyopathy | 161 |
| Distal myopathies | 38 | Limb girdle muscular dystrophy | 65 |
| Early onset dementia | 62 | Pulmonary arterial hypertension | 9 |
| Early onset Parkinson's disease | 172 | Rhabdomyolysis and metabolic muscle disorders | 27 |
| Epidermolysis bullosa | 16 |  |  |

PAX8 binding sites were identified using PAX8 ChIPseq data from eight cell lines: three benign immortalized normal fallopian tube cell lines (FT33, FT194 and FT246)^15^ and five EOC cell lines (KURAMOCHI, OVSAHO and JHOS4 ^15^; GROV1 and HeyA8 ^16^). PAX8 ChIPseq NarrowPeak formatted files are publicly available from the GEO (https://www.ncbi.nlm.nih.gov/geo/query/acc.cgi?acc=GSE79893) for the KURAMOCHI, OVSAHO and JHOS4 cancer cell lines and the FT33, FT194, and FT246 benign immortalized fallopian tube cell lines. PAX8 ChIPseq peaks for IGROV1 and HeyA8, as well as the signal-enriched and normalized files for all cell lines, were obtained from the Women’s Cancer Program at the Samuel Oschin Comprehensive Cancer Institute^16^. The number of PAX8 peaks and their genomic coverage for each cell line is shown in Supplementary Table 2.

Not all transcription factor binding sites will be active in any given tissue. Acetylation of histone H3K27 is a marker of active promoters and candidate enhancers and colocalizes with regulatory sequences and gene activation^17^. H3K27ac ChIPseq data were available for five different high-grade serous ovarian cancers and two normal fallopian tube epithelium cell lines (FT33 and FT246) (https://www.ncbi.nlm.nih.gov/geo/query/acc.cgi?acc=GSE68104) ^18^. The number of H3K27ac ChIPseq peaks for each cell line is shown in Supplementary Table 3.

**Supplementary Table 2:**
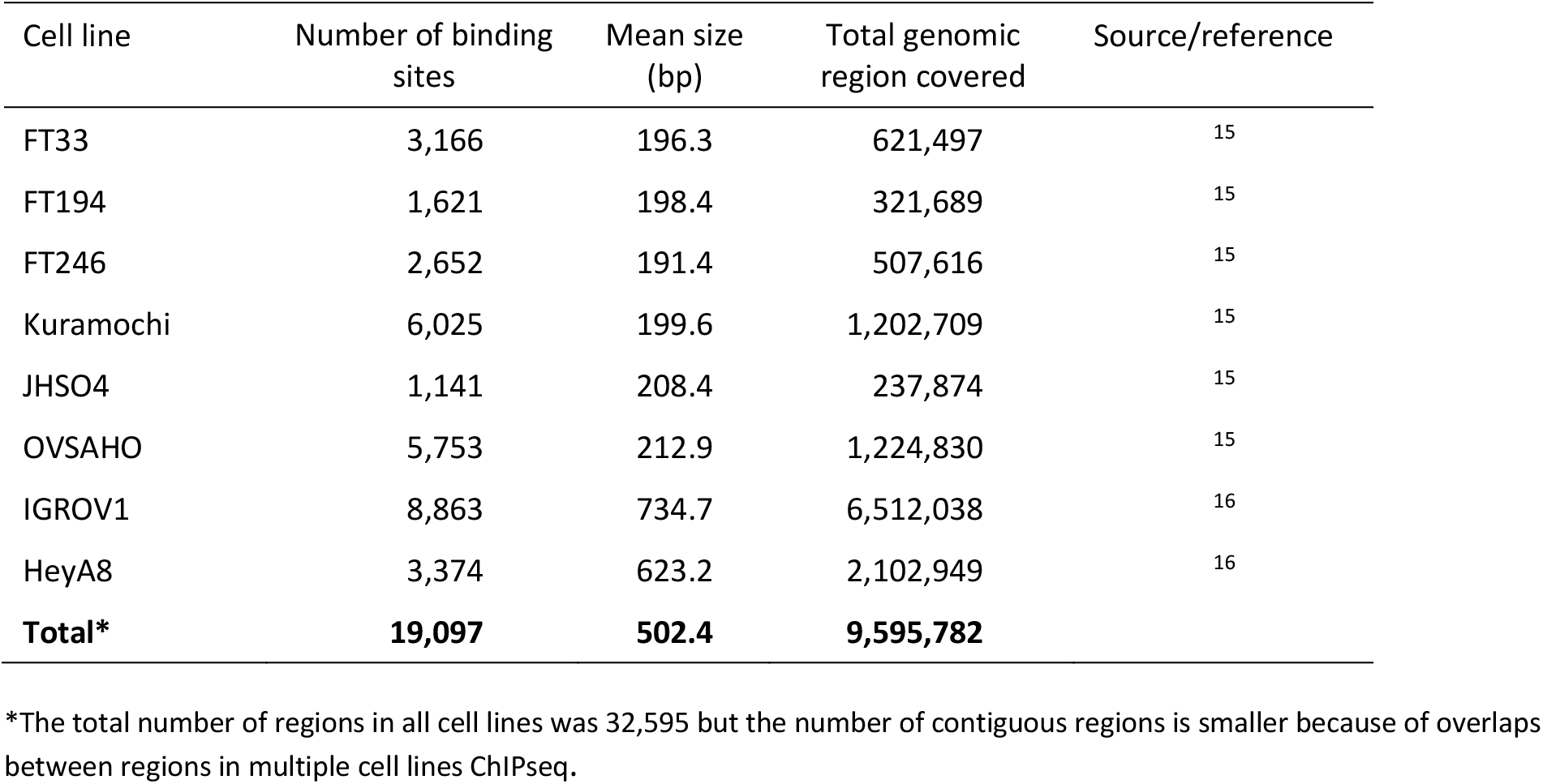
Number of PAX8 binding sites and genomic coverage for each of eight cell lines

**Supplementary Table 3:** Number of H3K27acetylation peaks and genomic coverage for each of the seven cell lines

| Cell line | Targets (n) | Mean size(bp) | Total coverage (bp) | Citation |
| --- | --- | --- | --- | --- |
| HGS_229 | 70,639 | 1314.0 | 92,822,771 | 19 |
| HGS_270 | 64,477 | 1415.9 | 91,294,827 | 19 |
| HGS_429 | 69,953 | 1049.3 | 73,405,375 | 19 |
| HGS_550 | 45,825 | 846.1 | 38,773,963 | 19 |
| HGS_561 | 75,628 | 1358.0 | 102,708,894 | 19 |
| FT33 | 74,475 | 1651.5 | 123,000,330 | 18 |
| FT246 | 86,534 | 1813.9 | 156,969,440 | 18 |
| Total | 164,268 | 1603.0 | 263,372,307 |  |

### Whole genome sequencing data

Germline whole genome samples were prepared using an Illumina TruSeq DNA Nano and sequenced at Illumina facilities using HiSeq X with 150 bases reads at a target of 30X in Genomics England facilities. Only samples with coverage of at least 95 percent of the genome at 15X or above with well mapped reads (mapping quality > 10) were included in the analyses. The Illumina North Star pipeline (version 2.6.53.23) was used for primary WGS analysis. Read alignment against human reference genome GRCh38-Decoy+EBV was performed with ISAAC (version iSAAC-03.16.02.19)^20, 21^

### Variant calling and quality control

All samples were processed with GATK Haplotype Caller version 4.0, with the standard thresholds for variant calling^22^. Variants were filtered out according to several quality criteria: read Depth <= 10; Mapping quality <= 20; allele balance <= 0.2 (proportion of alternate alleles) and alternative reads <=5; all variants in regions of low complexity. Low-complexity regions were identified using RepeatMasker^23^ and blacklisted regions from ENCODE^24^, resulting in the removal of 289,837 regions overlapping our target regions. Additional criteria were then used for exclusion of specific indels and single nucleotide variants (Supplementary Table 4).

**Supplementary Table 4:** Specific criteria for indel and single nucleotide variant exclusion

|  | Metric | Depth |  |  | Allele balance<br>>0.75 |
| --- | --- | --- | --- | --- | --- |
|  |  | > 100 | 60 - 100 | <60 |  |
| Indels | QD | <2.0 | <3.5 | <4.0 | <25 |
|  | MQ | <25 | <30 | <35 |  |
|  | FS | >50 | >45 | >40 |  |
|  | RPRS | <-5 | <-4.25 | <-3.5 |  |
| Single nucleotide variants | QD | <1.8 | <3.5 | <4.0 | <25 |
|  | MQ | <30 | <32.5 | <35 |  |
|  | FS | >50 | >45 | >40 |  |
|  | RPRS | <-5.5 | <-4.5 | <-3.5 |  |

QD = quality by depth; MQ = Mapping Quality ; FS = Phred-scaled p-value using Fisher's exact test to detect strand bias, RPRS = Z-score from Wilcoxon rank sum test of Alt vs. Ref read position bias.

These thresholds were based on an extensive comparison of concordance of whole exome sequence NGS calls with equivalent ChIP genotypes (unpublished). The rare variants were defined as those frequency lower than 0.01 in non-Finnish European samples or variants not reported in the gnomaAD 3.0 database^25^.

### Sample quality control

Sample quality control was carried out using the variants falling within the H3K_tum regions (defined in results section). The number of variants carried by each individual in these sets of regions varied from about 4,000 to over 20,000 with a long tail distribution (Supplementary Figure 1). We excluded 1,318 individuals that carried more than 6,800 rare alleles in the H3K_tum regions (Supplementary Table 5).

**Supplementary Figure 1:**
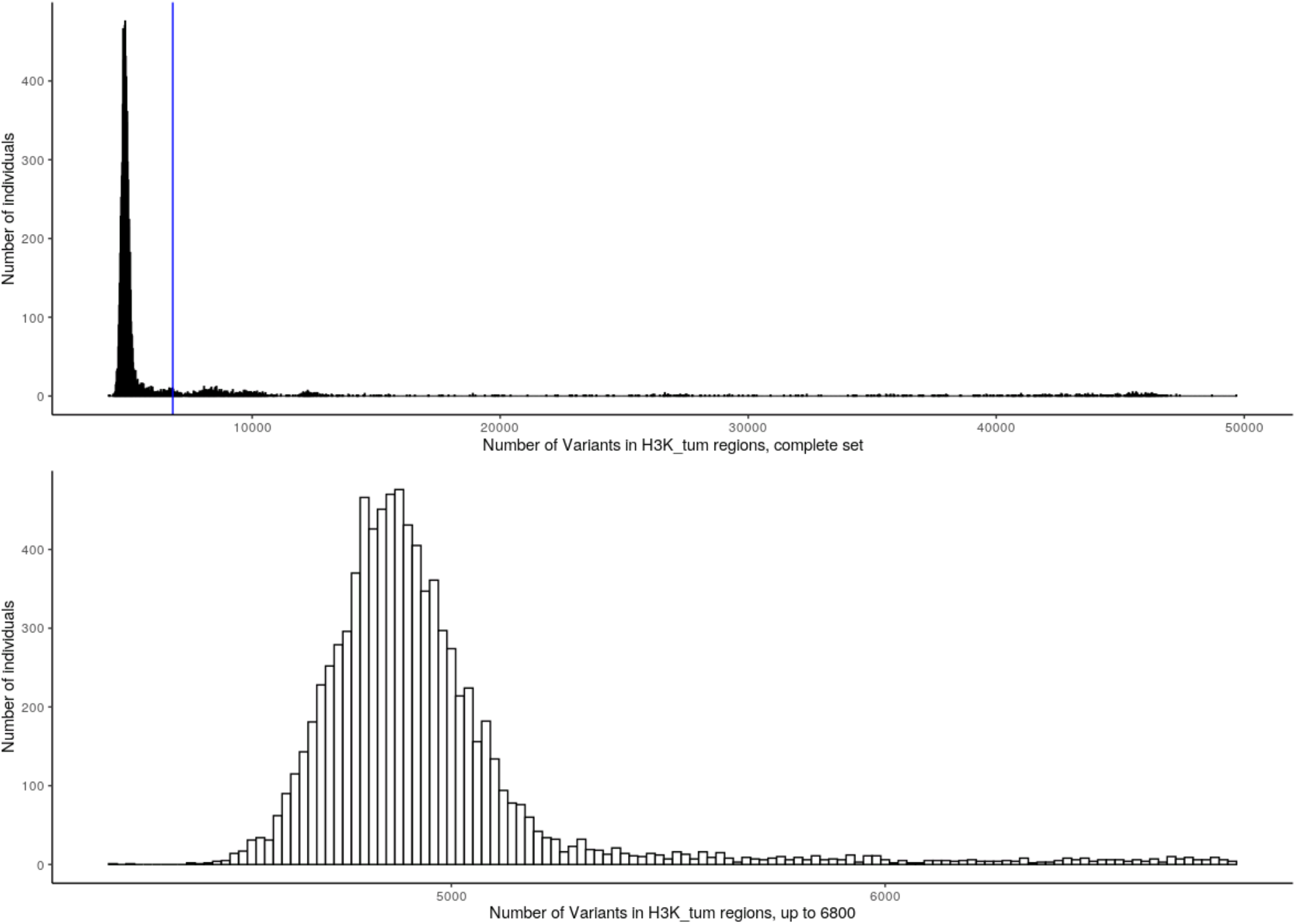
Histogram of the number of variants in H3K_tum regions in all patients.

The first plot represents the total number of variants found in all individuals; the blue line is the threshold admitted to proceed with further analysis. The second plot provides further details in the distribution of variants in H3K_tum regions, in all individuals.

**Supplementary Table 5:** Outlier patients removed (>6800 variants in H3K_tum regions)

| Samples | Outliers | Included (%) |
| --- | --- | --- |
| Control | 497 | 2253 (82) |
| Colorectal | 231 | 2465 (91) |
| Breast | 424 | 2560 (86) |
| Endometrial | 107 | 729 (87) |
| Ovarian | 59 | 522 (89) |

### Statistical methods

The number of variants in a given target region was compared across controls and the four cancers phenotypes using the non-parametric Kruskal-Wallis test. Then a pairwise comparison of each cancer phenotype with controls was carried out. We did not correct for multiple testing as the individual hypothesis tests are correlated, but given a low prior on the alternative hypothesis it is likely that most associations at a nominal P<0.05 would be false positives and so we set a nominal threshold for statistical significance at P<10^−3^.

## Results

### Defining the genomic region of interest

Across all the cell lines there were 19,097 peaks covering 9.5MB genomic DNA hereafter referred to as PAX8_all with PAX8_ft and PAX8_tum referring to PAX8 ChIPseq peaks in the fallopian tube and tumour cell lines respectively (Table 2). The number of H3K27ac ChIPseq peaks in the fallopian tube (H3K_ft) and tumours (H3K_tum) are shown in Table 2 with the numbers for each cell line individually in Supplementary Table 3.

**Table 2:** Selected genomic regions

| Region | Contiguous regions | Mean size | Total size (bp) |
| --- | --- | --- | --- |
| *PAX8 ChIPseq peaks* |  |  |  |
| All cell lines (PAX8_all) | 19,097 | 502.4 | 9,595,782 |
| Fallopian tube cell lines (PAX8_ft) | 4,468 | 199.2 | 890,331 |
| Tumour cell lines (PAX8_tum) | 17,772 | 525.2 | 9,333,999 |
| *H3K27ac ChIPseq peaks* |  |  |  |
| All cell lines | 164,268 | 1603.3 | 263,372,307 |
| Tumour cell lines (H3K_tum) | 129,293 | 1339.1 | 173,141,434 |
| Fallopian tube (H3K_ft) | 92,298 | 1813.3 | 167,368,077 |
| *Medium stringency regions* |  |  |  |
| All cell lines (MS_all) | 13,614 | 482.1 | 6,563,955 |
| Fallopian tube cell lines (MS_ft) | 2,672 | 187.3 | 500,604 |
| Tumour cell lines (MS_tum) | 11,585 | 493.7 | 5,719,946 |
| *High stringency regions* |  |  |  |
| All cell lines (HS_all) | 1,687 | 182.8 | 308,523 |
| Fallopian tube cell lines (HS_ft) | 2,701 | 191.1 | 516,210 |
| Tumour cell lines (HS_tum) | 947 | 180.1 | 170,628 |
| *Random regions* |  |  |  |
| Random 2K | 1,687 | 182.8 | 308,523 |
| Random 50k | 1,685 | 182.9 | 308,318 |
| Random Shuffle | 1,677 | 197.8 | 331,807 |
| Random H3K_tum | 113,888 | 987.7 | 112,494,386 |

We defined target regions based on the degree of overlap between PAX8 ChIPseq peaks and H3K27ac peaks, as can be observed in Figure 1. Medium stringency regions (MS_all) were those where there was overlap between H3K27ac marks in at least one of the seven cell lines (tumour or normal fallopian tubes) and PAX8 marks in at least one of the eight cell lines (tumour or normal fallopian tubes). Medium stringency fallopian tube regions (MS_ft) were those where there was overlap between H3K27ac marks in at least one of the two normal fallopian tube cell lines and PAX8 marks at least one of the three normal fallopian tube cell lines. Similarly, medium stringency tumour regions (MS_tum) were defined as those where there was overlap between H3K27ac marks in at least one of the five tumour cell lines and PAX8 marks in at least one of the five tumour cell lines. High stringency regions (HS_all) were defined as those where there was overlap between H3K27ac marks in at least three of the seven tumour or normal fallopian tube cell lines and PAX8 marks in at least three of the eight tumour or normal fallopian tube cell lines. High stringency fallopian tube regions (HS_ft) were those with at least three overlaps among any of the fallopian tube ChIPseqs. High stringency tumour regions (HS_tum) were those with overlap between H3K27ac marks in at least three of the five tumour cell lines and PAX8 marks in at least three of the five tumour cell lines. Following our prior experience, high stringency regions selected in this manner provide more robust evidence of signal^9^.

**Figure 1:**
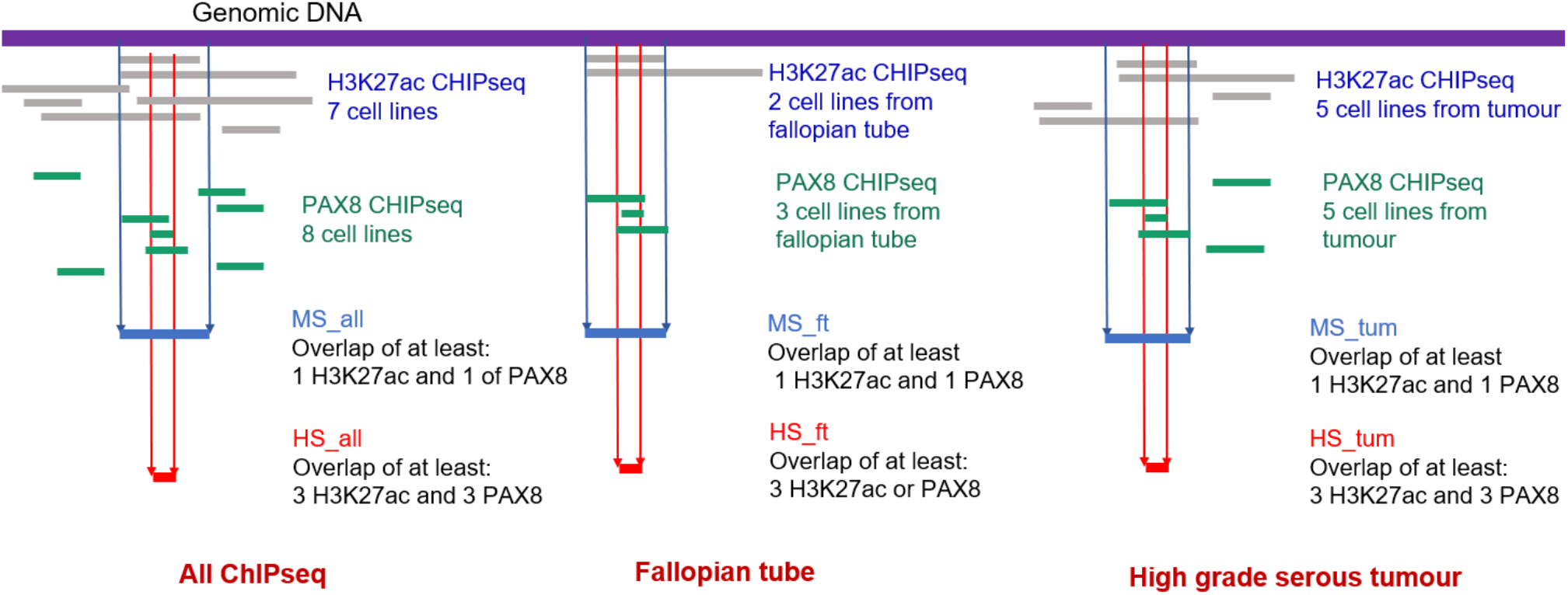
Schematic representation of the genomic regions of interest.

### Random control regions

Differential processing of the case and control samples from DNA extraction up to the sequencing and quality filters may result in artefactual differences in variant frequencies between cases and controls. In order to counteract the possible inflation of variants in the target regions, several types of control regions were created from random genomic sequences with similar characteristics to match the functional regions of interest. Variant frequencies in these random regions are unlikely to differ between cases and controls in the absence of functional elements.

Random sequences were selected to match 1,687 HS_all regions where there was overlap between 3 or more H3K27ac ChIPseq marks and 3 or more PAX8 ChIPseq marks. The criteria for a random sequence were that it must: i) be on the same chromosome ii) be the same size iii) similar base content with the ratio of the proportion of each base in the random region compared to the functional region being between 0.7 and 1.3 iv) not collocate with repeatMasker^23^ or ENCODE blacklisted regions^24^. Three types of random sequences were then chosen to match the HS_all regions: For the Random2k regions, a genomic window, the size of each of the 1,687 target regions was evaluated, starting at 2,000 bases upstream of the original site and moving 10bp upstream until the matching criteria was achieved. The Random50k regions were similarly selected starting at 50,000 bases upstream of the original site. Finally, we used *bedtools shuffle* ^26^ and seven different seeds to generate 8,620 regions from which 1,677 were selected to meet the criteria above (RandomShuffle regions).

A fourth type of random region was also selected to match the H3K_tum regions, using the same criteria as above and the same seven seeds for the random algorithm in *bedtools shuffle*, but the selection was a bit more stringent to yield a similar number of regions. The proportion was set between 0.75 and 1.25. This way, Random H3K_tum yielded 113,888 regions from the total of 129,293 H3K_tum target regions.

### Association of rare variants in PAX8 binding sites with cancer risk

The number of variants per sample by variant type (all variants, indels and substitutions) and phenotype in each of the eight defined functional regions are shown in Figures 2-4. There was nominally significant heterogeneity in the variant frequency across the controls and four case sets for the H3K_tum, H3K_ft, MS_all and MS_tum regions (Table 3). Pairwise comparisons for each set of cases and controls showed that this heterogeneity was being driven by a difference between colorectal cancer cases and controls.

**Table 3:** Kruskal-Wallis P-value for comparison of number of variants in cancer cases compared with controls (all variants)

| Region | All cases | Colorectal | Endometrial | Breast | Ovarian |
| --- | --- | --- | --- | --- | --- |
| H3K_ft | $1.9 \times 10^{-09}$ | $3.04 \times 10^{-08}$ | 0.92 | 0.28 | 0.22 |
| H3K_tum | $2.7 \times 10^{-9}$ | $1.2 \times 10^{-7}$ | 0.64 | 0.49 | 0.18 |
| PAX8 ChIPseq | 0.10 | 0.041 | 0.81 | 0.047 | 0.94 |
| MS_all | $2.3 \times 10^{-4}$ | $9.9 \times 10^{-5}$ | 0.93 | 0.050 | 0.48 |
| HS_all | 0.74 | 0.71 | 0.87 | 0.42 | 0.47 |
| MS_ft | 0.97 | 0.95 | 0.49 | 0.98 | 0.98 |
| HS_ft | 0.18 | 0.078 | 0.93 | 0.77 | 0.28 |
| MS_tum | $2.6 \times 10^{-4}$ | $1.1 \times 10^{-4}$ | 0.54 | 0.068 | 0.32 |
| HS_tum | 0.82 | 0.32 | 0.51 | 0.96 | 0.67 |
| Random2k | 0.022 | 0.42 | 0.21 | 0.42 | 0.013 |
| Random50k | 0.15 | 0.15 | 0.23 | 0.18 | 0.78 |
| RandomShuffle | 0.57 | 0.22 | 0.18 | 0.35 | 0.92 |
| RandomH3K_tum | $5.2 \times 10^{-9}$ | $5.2 \times 10^{-8}$ | 0.34 | 0.051 | 0.78 |

**Figure 2:**
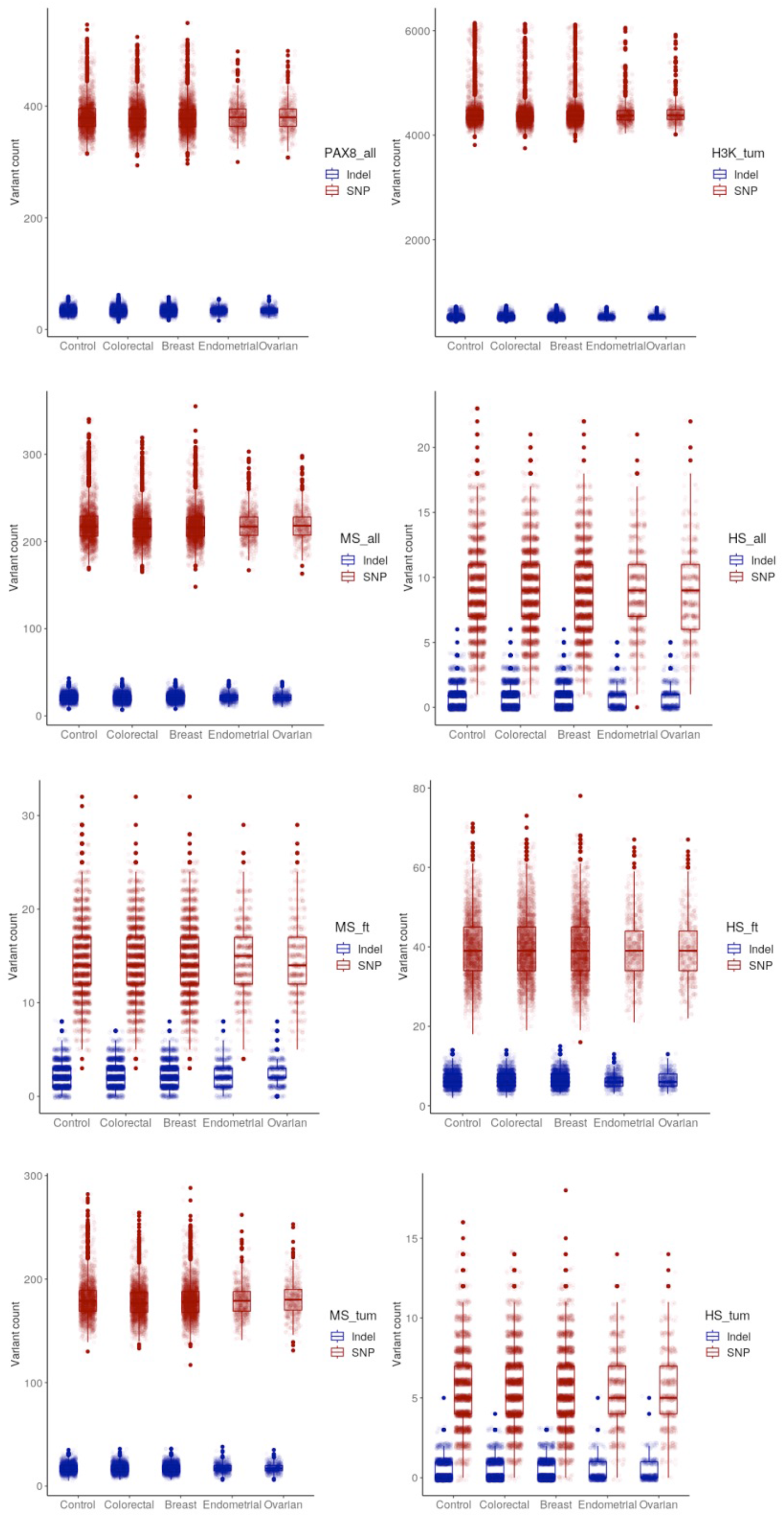
Number of variants per individual by case-control status and defined genomic region.

However, the observed differences were small and similar heterogeneity was seen for the Random H3K_tum regions (Supplementary Figure 2). This suggests that the observed differences are due to sequencing batch effects or similar technical artefact rather than any biological difference between cases and controls. When restricting the comparison to single nucleotide variants (Table 4) a similar pattern was seen. There were no nominally significant differences in the frequencies of indels in the target functional regions (Table 5).

**Table 4:**
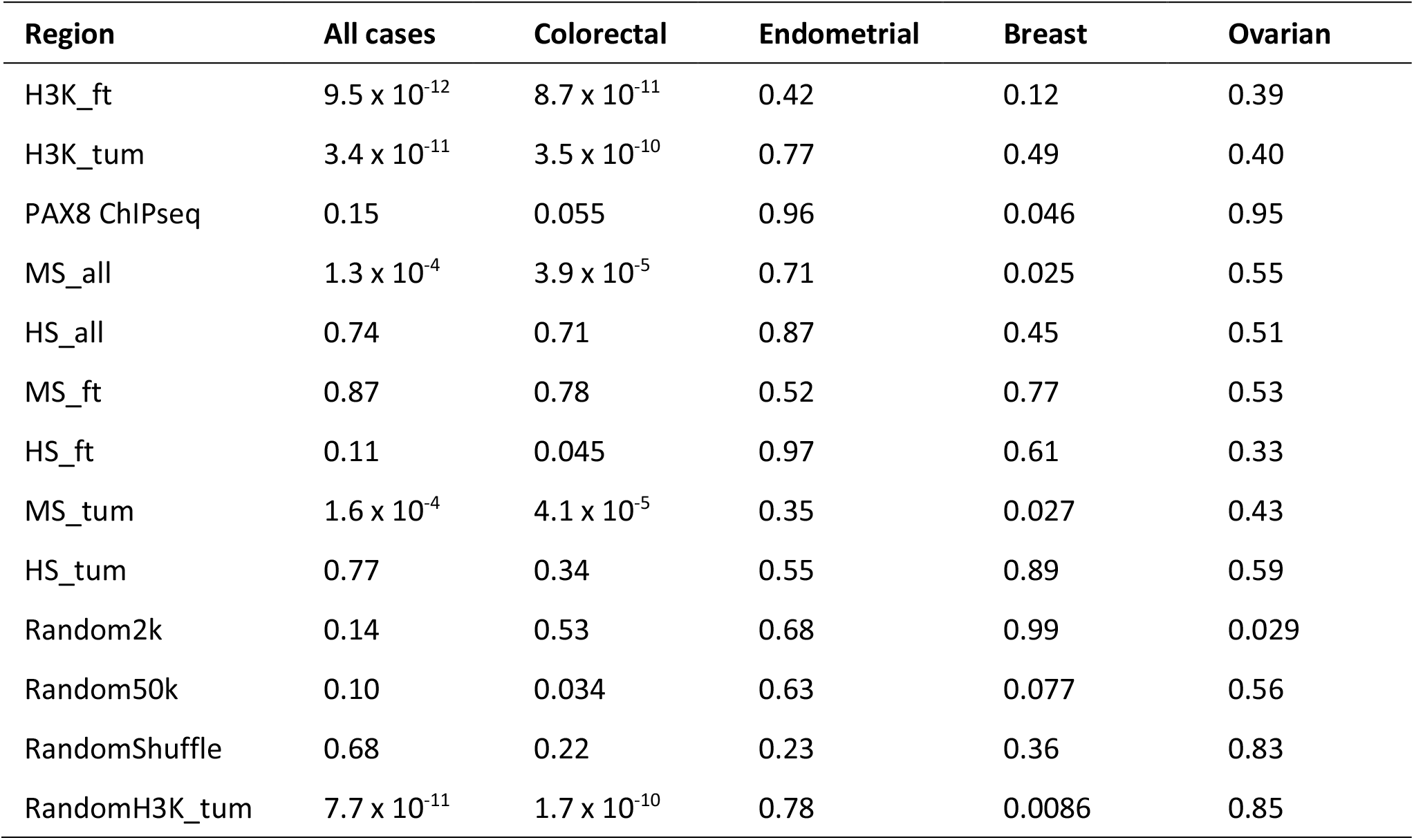
Comparison of number of variants in cancer cases compared with controls (Kruskal-Wallis P-value), only single nucleotide variation

| Region | All cases | Colorectal | Endometrial | Breast | Ovarian |
| --- | --- | --- | --- | --- | --- |
| H3K_ft | $9.5 \times 10^{-12}$ | $8.7 \times 10^{-11}$ | 0.42 | 0.12 | 0.39 |
| H3K_tum | $3.4 \times 10^{-11}$ | $3.5 \times 10^{-10}$ | 0.77 | 0.49 | 0.40 |
| PAX8 ChIPseq | 0.15 | 0.055 | 0.96 | 0.046 | 0.95 |
| MS_all | $1.3 \times 10^{-4}$ | $3.9 \times 10^{-5}$ | 0.71 | 0.025 | 0.55 |
| HS_all | 0.74 | 0.71 | 0.87 | 0.45 | 0.51 |
| MS_ft | 0.87 | 0.78 | 0.52 | 0.77 | 0.53 |
| HS_ft | 0.11 | 0.045 | 0.97 | 0.61 | 0.33 |
| MS_tum | $1.6 \times 10^{-4}$ | $4.1 \times 10^{-5}$ | 0.35 | 0.027 | 0.43 |
| HS_tum | 0.77 | 0.34 | 0.55 | 0.89 | 0.59 |
| Random2k | 0.14 | 0.53 | 0.68 | 0.99 | 0.029 |
| Random50k | 0.10 | 0.034 | 0.63 | 0.077 | 0.56 |
| RandomShuffle | 0.68 | 0.22 | 0.23 | 0.36 | 0.83 |
| RandomH3K_tum | $7.7 \times 10^{-11}$ | $1.7 \times 10^{-10}$ | 0.78 | 0.0086 | 0.85 |

**Table 5:** Comparison of number of indels in cancer cases compared with controls (Kruskal-Wallis P-value)

| Region | All cases | Colorectal | Endometrial | Breast | Ovarian |
| --- | --- | --- | --- | --- | --- |
| H3K_ft | $6.2 \times 10^{-03}$ | 0.66 | $4.4 \times 10^{-03}$ | 0.57 | 0.026 |
| H3K_tum | 0.014 | 0.80 | 0.011 | 0.16 | 0.0096 |
| PAX8 ChIPseq | 0.21 | 0.066 | 0.45 | 0.36 | 0.94 |
| MS_all | 0.47 | 0.14 | 0.87 | 0.65 | 0.64 |
| HS_all | 0.66 | 0.84 | 0.14 | 0.96 | 0.78 |
| MS_ft | 0.49 | 0.38 | 0.87 | 0.40 | 0.079 |
| HS_ft | 0.94 | 0.69 | 0.49 | 0.90 | 0.88 |
| MS_tum | 0.23 | 0.10 | 0.50 | 0.72 | 0.58 |
| HS_tum | 0.92 | 0.86 | 0.85 | 0.77 | 0.47 |
| Random2k | 0.0088 | 0.50 | 0.04 | 0.035 | 0.085 |
| Random50k | 0.63 | 0.59 | 0.12 | 0.58 | 0.98 |
| RandomShuffle | 0.38 | 0.76 | 0.32 | 0.47 | 0.18 |
| RandomH3K_tum | $8.9 \times 10^{-4}$ | 0.77 | $3.8 \times 10^{-4}$ | 0.10 | 0.0087 |

**Supplementary Figure 2:**
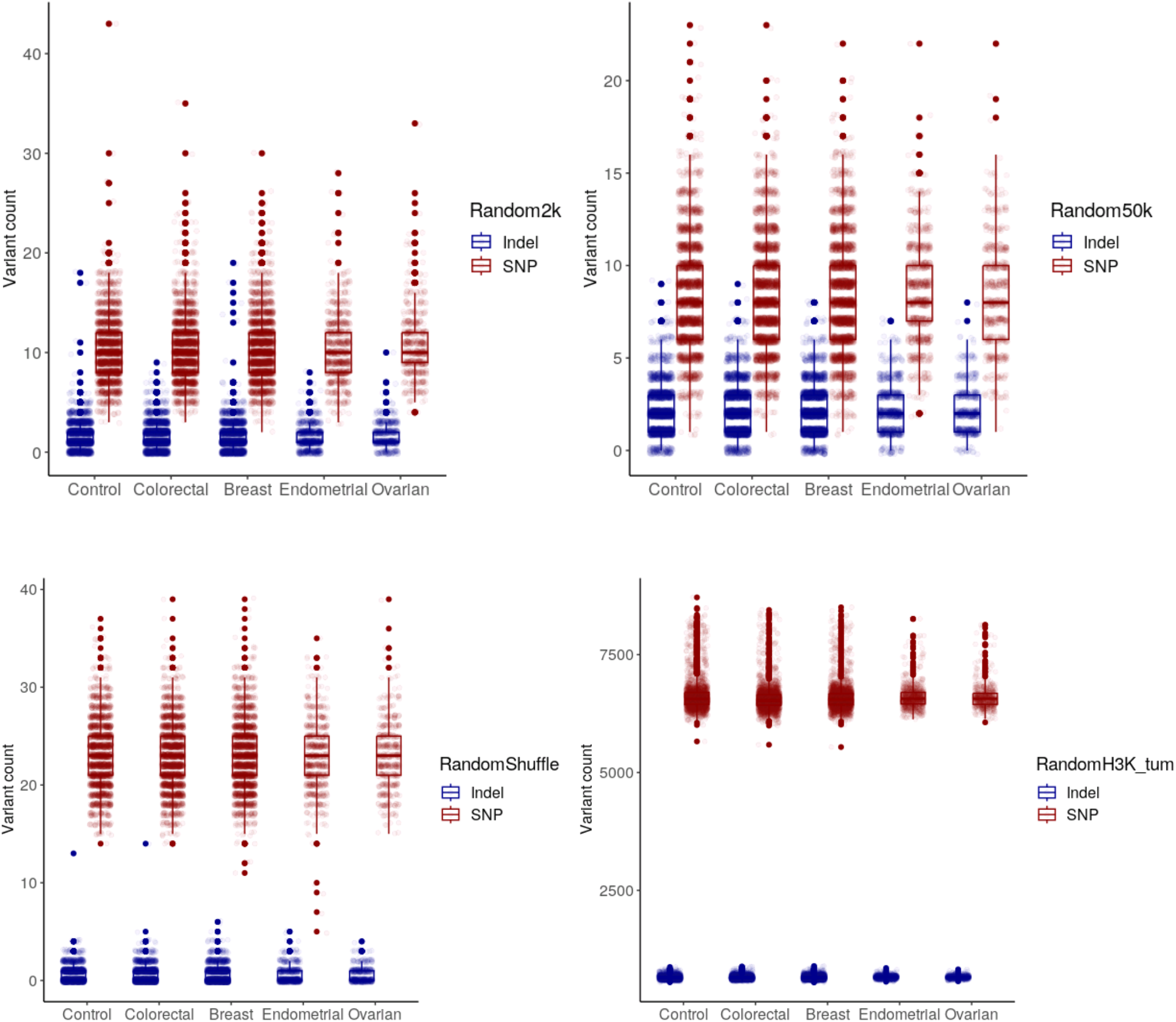
Number of variants per individual in Random Regions.

## Discussion

Rare, germline genetic variation in the non-coding genome is likely to account for a substantial portion of the unexplained heritability of many complex traits including ovarian cancer. Chip genotyping data combined with imputation can be used to evaluate rare variant associations, however, many rare variants are poorly imputed and whole genome sequencing is the only method to reliably capture all the rare variation across the genome. The evaluation of association for rare, non-coding variation on a variant-by-variant basis is unlikely to be an effective approach because of the very large number of such variants and the limited statistical power to detect association at very stringent levels of statistical significance required when the prior probability of any one association is very small. As with analysis of rare variation in the coding sequence, burden testing is an alternative approach to increase statistical power. However, unlike with gene-based burden testing, the best way to define groups of variants in the non-coding genome is not clear. One approach might be to simply combine rare variants within a specific genomic window. However, it seems very unlikely that multiple rare variants in any given contiguous genomic window would be functionally equivalent and associated with similar risks. We used an alternative approach, evaluating the burden variants in non-contiguous genomic regions that are thought to share some common function.

Many of the known common risk variants in the non-coding regions are thought to act by affecting regulatory elements. For example, breast cancer risk variants are enriched for binding sites of ESR1, FOXA1, GATA3, E2F1 and TCF7L2^27^ and six networks centred on homeobox transcription factor genes at 17q21.32 and at 2q31 are enriched for EOC susceptibility^28^. Thus, binding sites for transcription factors thought to be important in ovarian cancer biology would be good candidates for non-contiguous regions of the genome that harbour rare susceptibility variants for EOC.

We used publicly-available, germline, whole genome sequencing data of cancer and rare diseases patients to evaluate a hypothesised association between rare variation in putative active PAX8 transcription factor binding sites and risk of epithelial ovarian cancer. Our novel approach of using burden testing across non-contiguous regions of the genome has the potential to identify uncommon and rare germline genetic variants that underpin complex disease susceptibility. We found little evidence to support the hypothesis, but several limitations should be considered before considering these results definitive. First, statistical power is likely to be a limiting factor as there were data for only 522 EOC cases. A second limitation is our ability to identify PAX8 binding sites that are truly functional in the relevant tissue. We have used a variety of epigenomic data in an attempt to identify genomic regions that are most likely to be functionally relevant and we have used several different definitions to define putative functional regions. Nevertheless, it is likely that for any given definition of functionally relevant regions, a proportion of the binding sites of interest are not functional. It is also not fully understood how much a given variant would affect PAX8 binding if at all. Finally, redundancy of binding sites in the pathways activated by PAX8^9,10^ may limit the functional effect of a variant that does disrupt PAX8 binding in a relevant tissue. This misclassification could reduce substantially the power to detect a true association.

Despite the null findings and the limitations of this study, the approach based on burden testing across non-contiguous regions of the genome remains promising. Much large-scale case-control data will become available as whole genome sequencing becomes cheaper and this will increase substantially the statistical power of this type of study. Furthermore, improved functional annotation of the non-coding genome will lead to a better definition of the likely regions of the genome that are biologically important together with a better understanding of which variants are most likely to disturb putative functionality.

## Data Availability

Germline genetic data used in this project are available through application to the Genomics England 100,000 Genomes Project. CHiPSeq data are publicly available as described in the methods.

https://www.genomicsengland.co.uk

## References

1. Kar SP, Berchuck A, Gayther SA, et al. Common Genetic Variation and Susceptibility to Ovarian Cancer: Current Insights and Future Directions. Cancer Epidemiology, Biomarkers & Prevention. 2018;27(4):395-404. doi:10.1158/1055-9965.EPI-17-0315

2. Jones MR, Kamara D, Karlan BY, Pharoah PDP, Gayther SA. Genetic epidemiology of ovarian cancer and prospects for polygenic risk prediction. Gynecol Oncol. 2017;147(3):705–713. doi:10.1016/j.ygyno.2017.10.001

3. Miki Y, Swensen J, Shattuck-Eidens D, et al. A Strong Candidate for the Breast and Ovarian Cancer Susceptibility Gene BRCA1. Science (1979). 1994;266(5182):66–71. doi:10.1126/SCIENCE.7545954

4. Wooster R, Bignell G, Lancaster J, et al. Identification of the breast cancer susceptibility gene BRCA2. Nature. 1995;378(6559):789–792. doi:10.1038/378789A0

5. Kuchenbaecker KB, Hopper JL, Barnes DR, et al. Risks of breast, ovarian, and contralateral breast cancer for BRCA1 and BRCA2 mutation carriers. JAMA - Journal of the American Medical Association. 2017;317(23):2402–2416. doi:10.1001/jama.2017.7112

6. Shuai Li et al. Cancer Risks Associated With BRCA1 and BRCA2 Pathogenic Variants. J Clin Oncol. 2022;49:1529–1541. doi:10.1200/JCO.21.02112

7. Pavanello M, Chan IHY, Ariff A, Pharoah PDP, Gayther SA, Ramus SJ. Rare germline genetic variants and the risks of epithelial ovarian cancer. Cancers (Basel). 2020;12(10):1–23. doi:10.3390/cancers12103046

8. Wainschtein P, Jain D, Zheng Z, et al. Assessing the contribution of rare variants to complex trait heritability from whole-genome sequence data. Nat Genet. 2022;54(3):263–273. doi:10.1038/s41588-021-00997-7

9. Jones MR, Peng PC, Coetzee SG, et al. Ovarian Cancer Risk Variants Are Enriched in Histotype-Specific Enhancers and Disrupt Transcription Factor Binding Sites. Am J Hum Genet. 2020;107(4):622–635. doi:10.1016/j.ajhg.2020.08.021

10. Kar SP, Adler E, Tyrer J, et al. Enrichment of putative PAX8 target genes at serous epithelial ovarian cancer susceptibility loci. Br J Cancer. 2017;116(4):524–535. doi:10.1038/bjc.2016.426

11. Adler E, Mhawech-Fauceglia P, Gayther SA, Lawrenson K. PAX8 expression in ovarian surface epithelial cells. Hum Pathol. 2015;46(7):948–956. doi:10.1016/j.humpath.2015.03.017

12. Lawrenson K, Fonseca MAS, Liu AY, et al. A Study of High-Grade Serous Ovarian Cancer Origins Implicates the SOX18 Transcription Factor in Tumor Development. Cell Rep. 2019;29(11):3726-3735.e4. doi:10.1016/j.celrep.2019.10.122

13. Roadmap Epigenomics Consortium, Kundaje A, Meuleman W, et al. Integrative analysis of 111 reference human epigenomes. Nature. 2015;518(7539):317–329. doi:10.1038/nature14248

14. Creyghton MP, Cheng AW, Welstead GG, et al. Histone H3K27ac separates active from poised enhancers and predicts developmental state. Proc Natl Acad Sci U S A. 2010;107(50):21931–21936. doi:10.1073/pnas.1016071107

15. Elias KM, Emori MM, Westerling T, et al. Epigenetic remodeling regulates transcriptional changes between ovarian cancer and benign precursors. JCI Insight. 2016;1(13). doi:10.1172/jci.insight.87988

16. Adler EK, Corona RI, Lee JM, et al. The PAX8 cistrome in epithelial ovarian cancer. Oncotarget. 2017;8(65):108316–108332. doi:10.18632/oncotarget.22718

17. Roh TY, Cuddapah S, Zhao K. Active chromatin domains are defined by acetylation islands revealed by genome-wide mapping. Genes Dev. 2005;19(5):542–552. doi:10.1101/gad.1272505

18. Coetzee SG, Shen HC, Hazelett DJ, et al. Cell-type-specific enrichment of risk-associated regulatory elements at ovarian cancer susceptibility loci. Hum Mol Genet. 2015;24(13):3595–3607. doi:10.1093/hmg/ddv101

19. Corona RI, Seo JH, Lin X, et al. Non-coding somatic mutations converge on the PAX8 pathway in ovarian cancer. Nat Commun. 2020;11(1):1–11. doi:10.1038/s41467-020-15951-0

20. New models and technologies for personalised medicine. 2018;29(Supplement 6):2018. doi:10.1093/annonc/mdy318

21. Alona Sosinsky. Genomics England - Technical Information Document - Version 1.9.Main.; 2018.

22. Poplin R, Ruano-Rubio, Valentin DePristo MA, Fennell TJ, et al. Scaling accurate genetic variant discovery to tens of thousands of samples. BioRxIV. Published online 2018:1-22. doi:doi.org/10.1101/201178

23. Smit A, Hubley R, Green P. RepeatMasker Open-4.0. Published 2013. www.repeatmasker.org

24. Amemiya HM, Kundaje A, Boyle AP. The ENCODE Blacklist: Identification of Problematic Regions of the Genome. Sci Rep. 2019;9(1):1–5. doi:10.1038/s41598-019-45839-z

25. Karczewski KJ, Francioli LC, Tiao G, et al. The mutational constraint spectrum quantified from variation in 141,456 humans. Nature. 2020;581(7809):434–443. doi:10.1038/s41586-020-2308-7

26. Quinlan AR, Hall IM. BEDTools: A flexible suite of utilities for comparing genomic features. Bioinformatics. 2010;26(6):841–842. doi:10.1093/bioinformatics/btq033

27. Michailidou K, Lindström S, Dennis J, et al. Association analysis identifies 65 new breast cancer risk loci. Nature. 2017;551(7678):92–94. doi:10.1038/nature24284

28. Kar SP, Tyrer JP, Li Q, et al. Network-based integration of GWAS and gene expression identifies a HOX-centric network associated with serous ovarian cancer risk. Cancer Epidemiology Biomarkers and Prevention. 2015;24(10):1574–1584. doi:10.1158/1055-9965.EPI-14-1270

